# Reverse Mendelian randomization separates causes from early proteomic biomarkers of glioma

**DOI:** 10.1101/2024.03.29.24305009

**Authors:** Lily J Andrews, Zak A Thornton, Jie Zheng, Jamie W Robinson, Gibran Hemani, Kathreena M Kurian

**Affiliations:** MRC Integrative Epidemiology Unit (IEU), Bristol Medical School, University of Bristol, Oakfield House, Oakfield Grove, Bristol, BS8 2BN, United Kingdom; Population Health Sciences, Bristol Medical School, University of Bristol, Oakfield House, Oakfield Grove, Bristol, BS8 2BN, United Kingdom; Cancer Research Integrative Cancer Epidemiology Programme, University of Bristol, Oakfield House, Oakfield Grove, Bristol, BS8 2BN, United Kingdom; Brain Tumour Research Centre, Bristol Medical School, University of Bristol, Bristol, UK; Department of Endocrine and Metabolic Diseases, Shanghai Institute of Endocrine and Metabolic Diseases, Ruijin Hospital, Shanghai Jiao Tong University School of Medicine, Shanghai, China; Shanghai National Clinical Research Center for metabolic Diseases, Key Laboratory for Endocrine and Metabolic Diseases of the National Health Commission of the PR China, Shanghai Key Laboratory for Endocrine Tumor, State Key Laboratory of Medical Genomics, Ruijin Hospital, Shanghai Jiao Tong University School of Medicine, Shanghai, China

## Abstract

**Background/Objectives:** Glioma represents the largest entity of primary brain tumours in adults, with an overall survival of less than 20% over 5 years. Glioblastoma is the most frequent and aggressive glioma subtype. At present, there are few well-established pre-clinical predictors for glioma incidence. Due to the availability and size of prognostic studies in glioma, we utilised a Mendelian randomization framework to identify non-causal protein biomarkers which are associated with early-onset of glioma in the European population.

**Methods:** We generated polygenic risk scores (PRS) for glioma (n=12,496), glioblastoma (n=6,191), and non-glioblastoma (n=5,819) cases. We used reverse Mendelian randomization (MR) to examine the relationship between the genetic liability of glioma and 1,463 and 90 proteins were measured using an Olink panel (UKBB, n=35,571 and SCALLOP, n=21,758), additionally 4,907 and 2,994 aptamers were assayed using SOMAscan assays (deCODE n=35,559 and INTERVAL, n=3,301). We further performed a forward cis-MR and colocalization analysis leveraging the circulating protein markers in risk of glioma, glioblastoma and non-glioblastoma.

**Results:** Reverse MR identified 161 unique proteins associated with the PRS of glioma, 79 proteins associated with the PRS of glioblastoma, and 11 proteins associated with the PRS of non-glioblastoma. Enrichment analyses identified a proportion of plasma proteins to be associated with the PRS of glioma to be correlated with response to external stimulus. A group of plasma proteins linked to the PRS of glioma and glioblastoma were related to the immune system process. Forward MR of the putative relationships were found to have little or no evidence of association on the causal pathway. Candidate markers ETFA, RIR1 and BT3A1 are evidenced in glioma risk.

**Conclusion:** Our findings identify a high genetic liability to glioma is associated with the immune system processes. Non-causal plasma biomarkers identified through PRS associations could indicate novel non-causal biomarkers of early glioma development.

## Background

Central nervous system (CNS) and brain tumours lead to significant years of life lost compared to other cancer types, with an average of over 20 years of life lost(1). Gliomas are the largest group of intrinsic brain tumours, with age-adjusted incidence rates ranging from 4.67 to 5.73 per 100,000(2). At present, the gold standard diagnostic tool to detect brain tumours are MRI scans(3). A blood-based liquid biopsy could provide a cheap, simple, and minimally invasive way to diagnose brain tumours and monitor for recurrence(4).

Glioblastomas are the most common and malignant glioma subtype, representing around 55% of gliomas. Glioblastoma tumours are highly invasive malignant tumours with a survival rate of less than 5% after 5 years(5). In the 2021 World Health Organisation (WHO) classification of CNS tumours, non-glioblastoma gliomas are suggested to be less malignant and sometimes characterised by an isocitrate dehydrogenase mutation; the most common entities include astrocytoma and oligodendroglioma(6). Identification of novel biomarkers has relied heavily on laboratory-based liquid biopsy proteomic studies using clinical cohorts and small protein panels. However, recent advances have meant large genome wide-association studies (GWAS) of the plasma proteome are cheap and feasible to conduct. This has led to GWAS identifying genetic variants associated with thousands of circulating proteins, called protein quantitative trait loci (pQTL)(7–12). Generally, pQTLs have been used to proxy the effects of perturbed protein abundance on disease risk to aid drug target identification(13), such as using forward Mendelian randomization (MR) which is a statistical method for causal inference which uses genetic variants to proxy the effects of an exposure, generally a modifiable risk factor, on an outcome, which is usually a phenotype or disease(14). However, recently a novel application of these data in a MR framework has been used to identify putative diagnostic biomarkers for the early detection of disease(15).

For disease prediction, we consider the liability threshold (LT), this refers to the tendency for an individual to develop disease based on both environmental and additive genetic factors(16). In a normally distributed model for disease, there is a point, known as the threshold, at which all individuals below are considered unaffected and everyone above that point are affected(17). Liability is a continuous lifetime exposure, and as mentioned earlier manifests into disease in a small subset of the population but likely has accumulating biological consequences on all individuals, relative to their liability. Protein levels that respond to such processes may indicate individuals with a higher disease liability.

To identify proteins related to disease liability, a novel approach conceived by Mohammadi-Shemirani et al, which they called “reverse MR”, this generates a PRS for disease liability and tests for associations with proteins levels in the population studied(15). The hypothesis behind the reverse MR framework is that the biological consequence of disease liability alters proteomic profiles of individuals pre-onset of disease, and these biomarkers can be useful tools for prediction and diagnostics(18). Proteins identified to be a consequence of disease liability will associate with all liability SNPs, and we hypothesize these will be identified in the reverse MR analysis. It is important to note that this evidence of association will likely not indicate proteins that are causal for disease liability, as contrary to a standard forward MR study certain assumptions which underpin the MR methodology are relaxed. Moreover, proteins measured in this two-sample reverse MR framework are from the population studied, e.g. Europeans, with whom almost all individuals (due to the rarity of the disease) are not diagnosed with glioma nor will not go on to develop it.

In this paper, we aim to investigate if reverse MR can identify non-causal protein predictors for the early detection of glioma, and which processes these proteins may be involved in. We also aim to identify putative plasma protein biomarkers involved in glioma risk using forward MR.

## Methods

### Ethics

Ethical approval is not required as this study is using previously published GWAS where ethical approval has already been granted.

### Overview

We weighted four polygenic risk scores from glioma, glioblastoma and non-glioblastoma entities and used genetically-proxied protein levels from four European cohorts to carry out the reverse MR. We used the same datasets, instead selecting instruments from the protein cohorts in the forward MR approach and carried out a colocalization analysis. For the MR methods we performed sensitivity analyses, and then a quantitative trait loci enrichment analysis. We also searched the literature for any pre-diagnostic protein biomarkers that associate with glioma entities which may provide more insight to our non-causal early markers found in the reverse MR.

### Polygenic risk score weights

The glioma dataset is derived from a meta-analysis of 8 different GWAS; UK-GWAS, French-GWAS, German-GWAS, MDA-GWAS, SFAGS, GliomaScan, GICC, and UCSF-Mayo. These studies total to 12,496 adult (aged >18 years) gliomas cases (6,191 glioblastoma and 5,819 non-glioblastoma) and 18,190 controls with European descent. Cases were newly diagnosed with glioma and the controls were recorded as having no history of central nervous system tumours at the time of extraction (Supplementary Table 1)(19).

We constructed four sets of polygenic risk score weights for glioma, glioblastoma and non-glioblastoma using the traditional clumping and threshold (C and T) approach. This means the data is restricted by GWAS P-value and LD clumping thresholds. Each set of PRS weights was labelled P1 to P4, where P1 represented the most lenient threshold to P4 which represented the most stringent threshold (Table 1). Two PRS weights were used in our main analysis (P1 and P4), to ensure the PRS was robust across different weights, the other two were included in the sensitivity analyses (P2 and P3).

**Table 1:** The four parameters to generate each polygenic risk score for all glioma cases, glioblastoma and non-glioblastoma.

| Parameter | P-value | $r^2$ |
| --- | --- | --- |
| One | $1.00 \times 10^{-5}$ | 0.1 |
| Two | $1.00 \times 10^{-5}$ | 0.01 |
| Three | $5.00 \times 10^{-8}$ | 0.1 |
| Four | $5.00 \times 10^{-8}$ | 0.01 |

### Plasma proteome cohorts

The UK Biobank cohort (UKBB) measured protein levels of 35,571 participants (between 40-69) of European ancestry using the Olink Explore 1536 platform(Supplementary Table 2) (20). The deCODE study measured 4,907 aptamers in 35,559 Icelanders using SomaScan version 4 assay, this is made up of the Icelandic Cancer Project (52%)(21) and deCODE genetics(Supplementary Table 2)(22). The SCALLOP study consists of 13 cohorts of European ancestry of 21,758 individuals with 90 unique proteins measured using the Olink PEA CVD-I panel(Supplementary Table 2)(23). In the INTERVAL study, summary statistics for 3,301 individuals aged 18 years or over and of European descent had 2,994 cis-acting and trans-acting plasma pQTLs measured by SomaScan assay(Supplementary Table 2)(8).

### Reverse Mendelian Randomization

To assess the associations between the genetic liability to glioma, glioblastoma, non-glioblastoma and genetically-inferred circulating protein levels, we used reverse Mendelian randomization (MR). Our hypothesis of this statistical test is that the PRS of disease gives an indication of disease liability (Figure 1). As we are using genetically proxied protein levels from European populations, the hypothesis is that reverse MR will identify non-causal markers which contributes to a high genetic liability in European individuals to glioma and that these markers may be better as diagnostic markers compared to causal markers.

**Figure 1:**
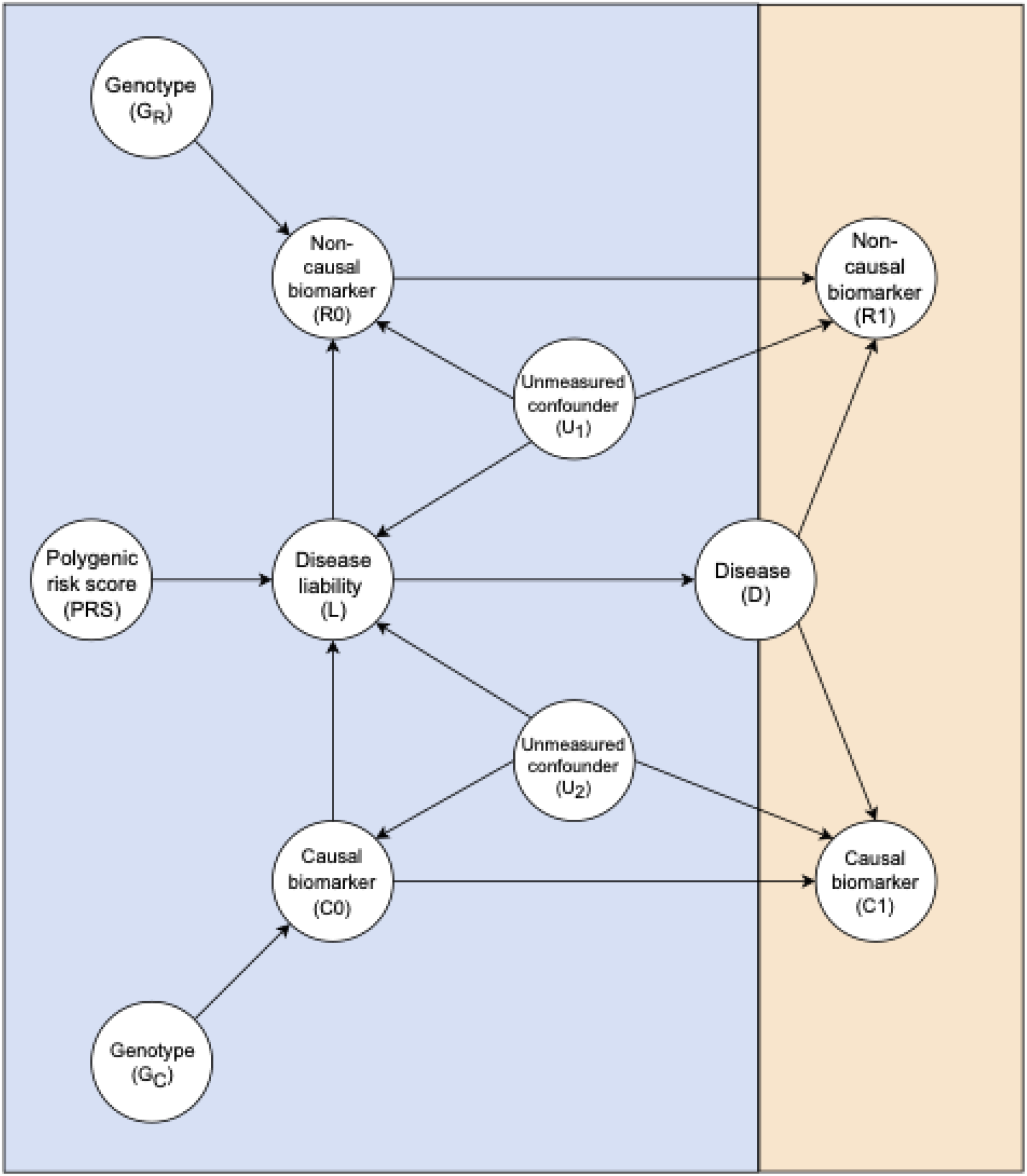
Direct acyclic graph representing hypothesis of reverse Mendelian randomization in disease liability.

To carry out the reverse MR, we leveraged the polygenic risk scores for glioma, glioblastoma and non-glioblastoma (using both cis-acting and trans-acting genetic variants, to increase the proportion of variance explained by the variants on the exposure) and the proteome cohorts listed above. We used the TwoSample MR package (24, 25) with PRS for glioma, glioblastoma and non-glioblastoma as the exposures, and the proteome cohort as the outcome dataset.

For the main analysis, we only looked at estimates generated by the inverse variance weighted method as all the exposures had multiple associated SNPs. For the P1 threshold, a false discovery rate (FDR) value was estimated for each protein to account for multiple testing. If an association from the P1 analysis passed q-value<0.05 and MR P-value<0.05 in the P4 analysis, this would be considered a strong association.

We also assessed heterogeneity between the SNPs in the reverse MR effect estimates for P1, for this we performed a Cochrane’s Q-test. If Cochrane’s Q-test<0.05, this result could be deemed as potential heterogeneity. To avoid reporting results in which a single SNP was driving the association, for associations with potential heterogeneity we carried out a leave-one-out analysis. If the leave-one-out analysis contained SNPs which differed by >0.01 this was deemed as a highly heterogenous association and was not included in our main analyses. We performed independent replication by comparing the similarity of the effect estimate between the UKBB cohort against deCODE, SCALLOP and INTERVAL protein associations (this included any proteins which reverse MR was carried out) using a pairwise correlation plot. We also compared the expected versus observed signs of beta values generated in the reverse MR result between UKBB (as the discovery beta) and deCODE (as the replication beta).

### Forward Mendelian randomization

We performed a two-sample forward MR analysis to estimate the causal effect between genetically proxied protein abundance and glioma risk in the same datasets as above. Hence in this analysis, proteome cohorts were the exposure datasets and the glioma, glioblastoma and non-glioblastoma datasets were the outcome datasets. Instruments were selected at a GWAS P-value threshold of <5.0×10^−8^ and clumped at a LD threshold of *r*^2^=0.001. pQTLs from the proteome cohorts were classified as cis-acting, which is defined as a SNP in the region of 1Mb window of the gene regulatory region, or trans-acting which is defined as a SNP located outside of the cis region. Only cis-acting SNPs were included in the two-sample forward MR analysis to mitigate the influence of horizontal pleiotropy in our results.

We carried out the same multiple testing on the results as used in the reverse MR which generated an FDR value for each protein. If an association had a q-value<0.05 it is considered a strong association, and if a q-value was between 0.1 and 0.05 this would be considered a suggestive association.

### Colocalization

For putative proteins with a q-value<0.1 (q-value<0.1 a suggestive association and q-value<0.05 a strong association) in the forward MR analysis, we carried out a pair-wise conditional and colocalization analysis using the PWCOCO package on Linux (26, 27). This involved selecting variants in a +/− 500kb window of the gene of interest for both the proteome and glioma dataset. Each of the five hypotheses are tested to see if both the exposure and outcome dataset has the same shared variant driving the association. A posterior probability (PP) is generated to suggest the relationship between variants. If the posterior probability of hypothesis 4 (PPH4) has the largest percentage, this proposes SNPs are colocalized and share the same variant. For this analysis, we considered PPH4>80% as strong evidence of colocalization, 80%≤PPH4≥50% as moderate evidence of colocalization, and PPH4<50% as weak evidence of colocalization.

### Reverse Mendelian randomization: sensitivity analyses

We compared the putative proteins discovered in the forward MR against the proteins identified in the reverse MR. This is due to our hypothesis that reverse MR is best powered to generate non-causal markers for disease and forward MR is well powered to generate causal markers for disease. Therefore, these analyses should not have the same proteins evidenced with disease risk.

We looked at the direction of effect for each association in MR-Egger and weighted median regression scores to further assess the validity of our findings. The two other PRS (P2 and P3) for glioma, glioblastoma and non-glioblastoma were also investigated as part of our sensitivity analysis, to see if these associations had an MR P-value<0.05. We also ensured each SNP included in the reverse MR analysis was not located in the MHC region on chromosome 6, due to the complex LD structure in this region.

### Forward Mendelian randomization: sensitivity analyses

For all molecular traits with a q-value<0.1 We carried out a Steiger filtering test to assess directionality of the causal association. This is to determine whether there could be reverse causation between the protein and glioma entity which is driving the associations identified. If the Steiger filtering test was “True” in the direction of exposure to outcome this would pass the test, so the association would be considered the correct direction. If the Steiger filtering test is “False”, this would suggest the direction of effect is reverse causal. Tests for heterogeneity and effect estimates were followed up but as the associations that passed q-value<0.01 had 5 or less SNPs, these tests were not as robust as the tests presented in the reverse MR analyses so these are reported in supplementary section 1.

### Statistical analysis

The reverse MR and forward MR analysis were supported by the IEU OpenGWAS database and ‘TwoSampleMR’ R package which was carried out on RStudio(24, 25, 28-30). FDR was calculated using ‘stats’ package in RStudio(29). Pairs and lm function on base R was used to generate a pairwise correlation plot comparing UKBB proteins with other cohorts(31). These packages were run using R version 4.0.3(31).

### Quantitative Trait Loci Enrichment Analysis

For associations discovered in the reverse MR analyses we searched STRINGdb for the enrichment of these proteins to identify if these were co-expressed, and which pathways may be enriched(32). Similarly, we carried this out for proteins identified in the forward MR analysis. An enrichment p-value is generated in STRINGdb, so we sampled the same number of proteins as identified in our main analysis for glioma, glioblastoma and non-glioblastoma from the combined cohorts (UKBB, deCODE, SCALLOP and INTERVAL). We generated these randomly selected proteins 100 times and compared these values to the enrichment value from the main analysis. We then generated an empirical p-value for the main analysis depending on the position this value was within the 100 sampled p-values. For the glioma entity analysis, the p-value is so low for the database it is considered as 0 in the API From the 100 generations of random proteins there were 5 scenarios where the enrichment p-value is 0, therefore we can’t decipher which p-value is lower between these and the enrichment from the main analysis.

### Literature Search

We carried out a literature search of any markers identified in our reverse MR analysis to see if any of these markers have been previously identified in nested case control studies. This includes protein measures before glioma diagnosis, also known as pre-diagnostic markers.

## Results

### Reverse MR

A total of 161 unique proteins were found to be associated with the glioma PRS, these proteins were made up of 70 in UKBB, 86 in deCODE, 5 in INTERVAL, and none were from the SCALLOP cohort (Supplementary Table 3-5). To note CAMP was present twice in deCODE, but these were two aptamers. There were 79 proteins which associated with the glioblastoma PRS, 35 in UKBB, 42 in deCODE, 2 in INTERVAL, and none were identified in SCALLOP (Supplementary Table 6-8). Finally, 13 proteins were associated with the non-glioblastoma PRS 11 in UKBB, 2 in SCALLOP and none identified in deCODE or INTERVAL cohorts (Supplementary Table 9-10).

Of these proteins discovered to be associated with glioma PRS, three proteins were removed for having potential high heterogeneity, these were EDA2R, FLT3LG and KLRB1 (Supplementary Table 11). These were originally UKBB proteins associated with glioma and glioblastoma PRS. Most of the associations were found to have a MR P-value<0.05 in P2 and P3 threshold, with the exception of 25 proteins associated with glioma PRS not passing P2 and 1 protein not passing P3. For glioblastoma, 17 proteins associated with glioblastoma PRS did not pass P2 MR P-value<0.05, but all passed P3.

For the UKBB proteins discovered in the reverse MR, we looked to see if the betas of these proteins followed a similar pattern within deCODE and INTERVAL cohorts. For glioma and glioblastoma, the cohorts follow the same effect pattern (Figure 2 and 3). We tested the concordance of beta signs between UKBB (as the discovery betas) and deCODE (as the replicated betas) were tested. Accounting for differences in power, we expected 39 out of 39 UKBB discovery associations to have the same sign in deCODE, however we only observed 29 to have the same sign. For the glioblastoma subset we observed 28 to have the same sign compared to an expected 31 and for the non-glioblastoma subset we observed 7 to have the same sign of an expected 8. Non-glioblastoma estimates did not seem to associate as well between INTERVAL and the other cohorts, but this could be due to the smaller number of proteins included in the analysis (Figure 4).

**Figure 2:**
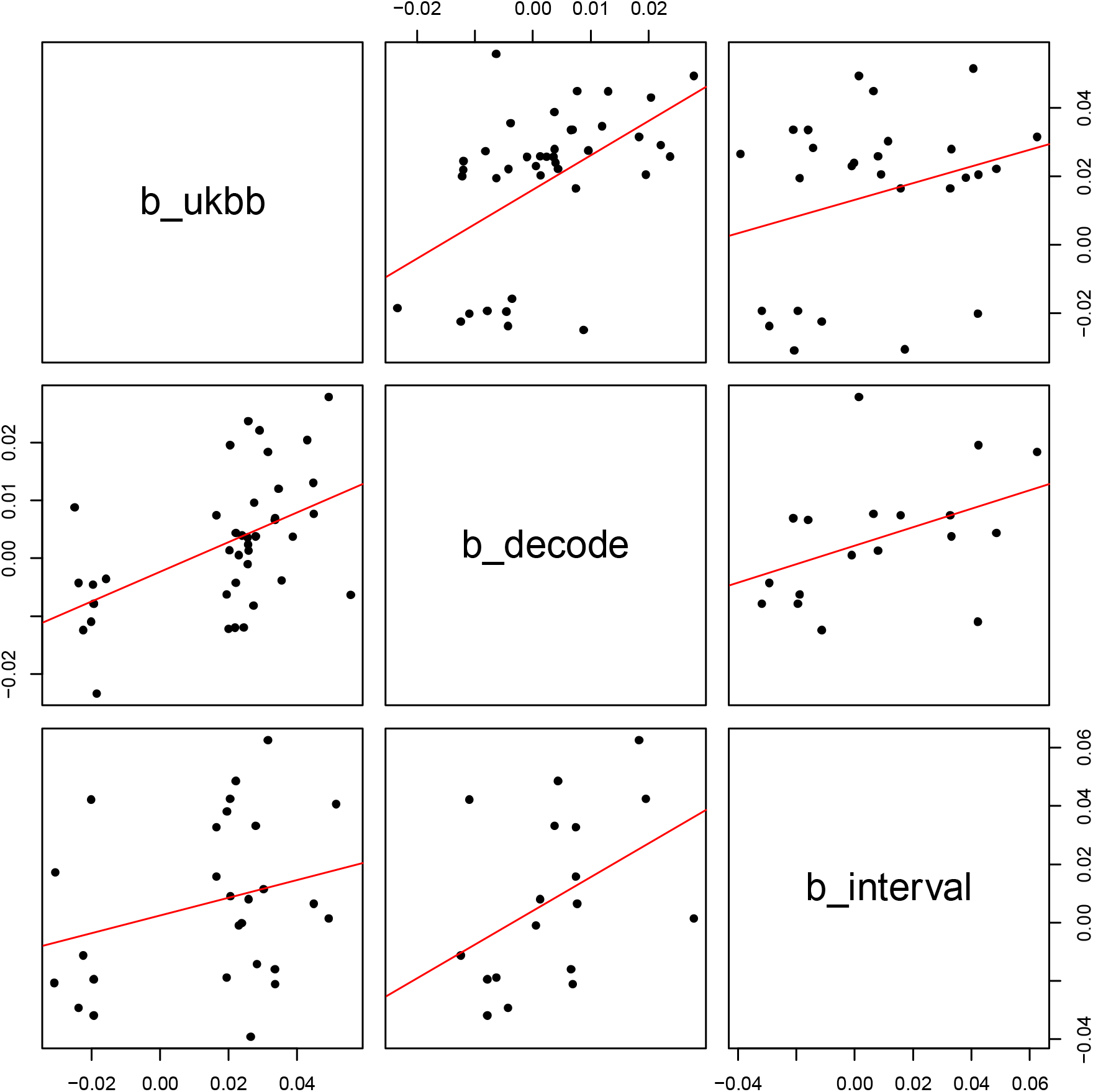
Proteins associated with glioma PRS in the UKBB cohort compared with these proteins in deCODE, INTERVAL and SCALLOP.

**Figure 3:**
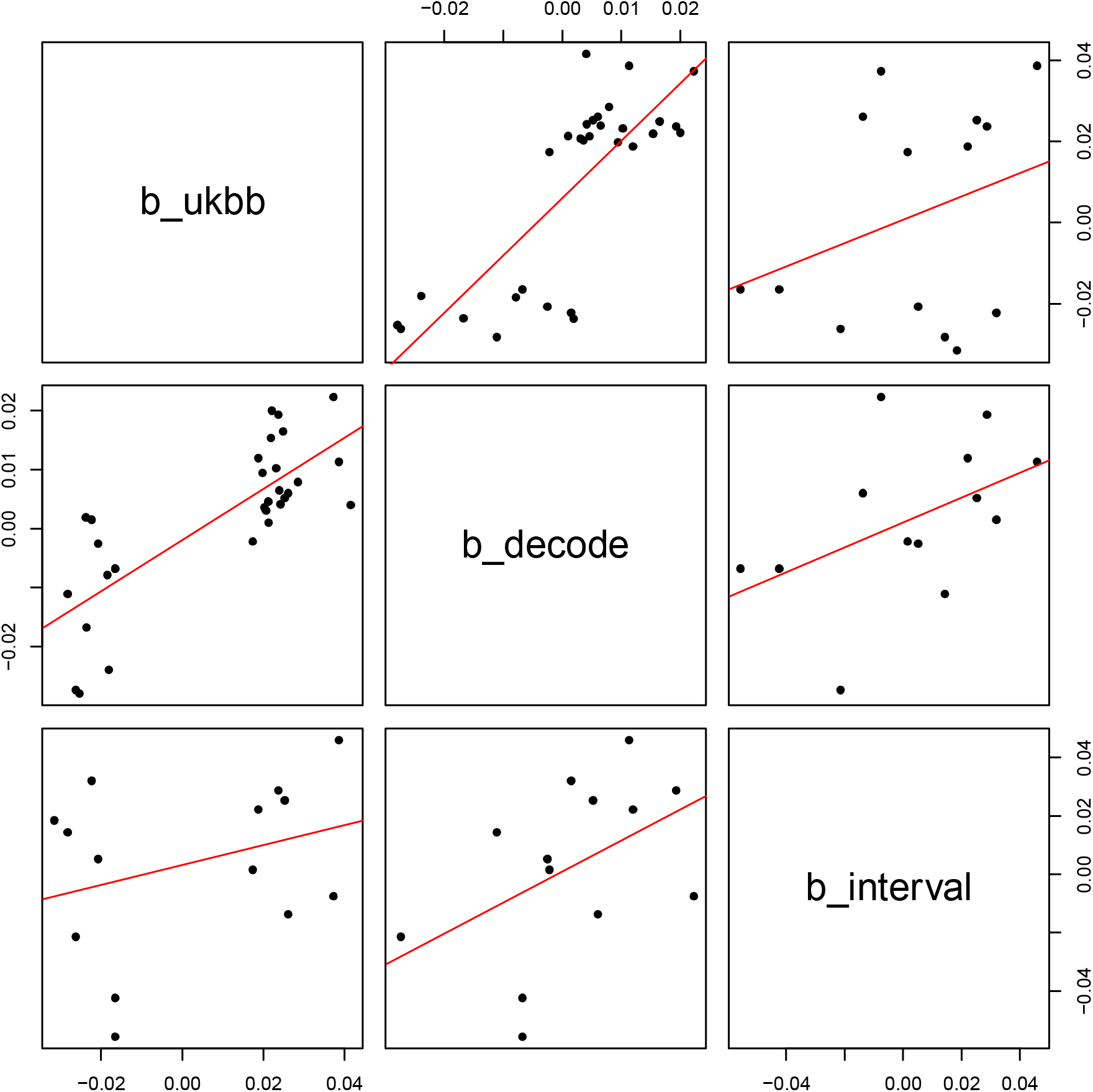
Proteins associated with glioblastoma PRS in the UKBB cohort compared with these proteins in deCODE, INTERVAL and SCALLOP.

**Figure 4:**
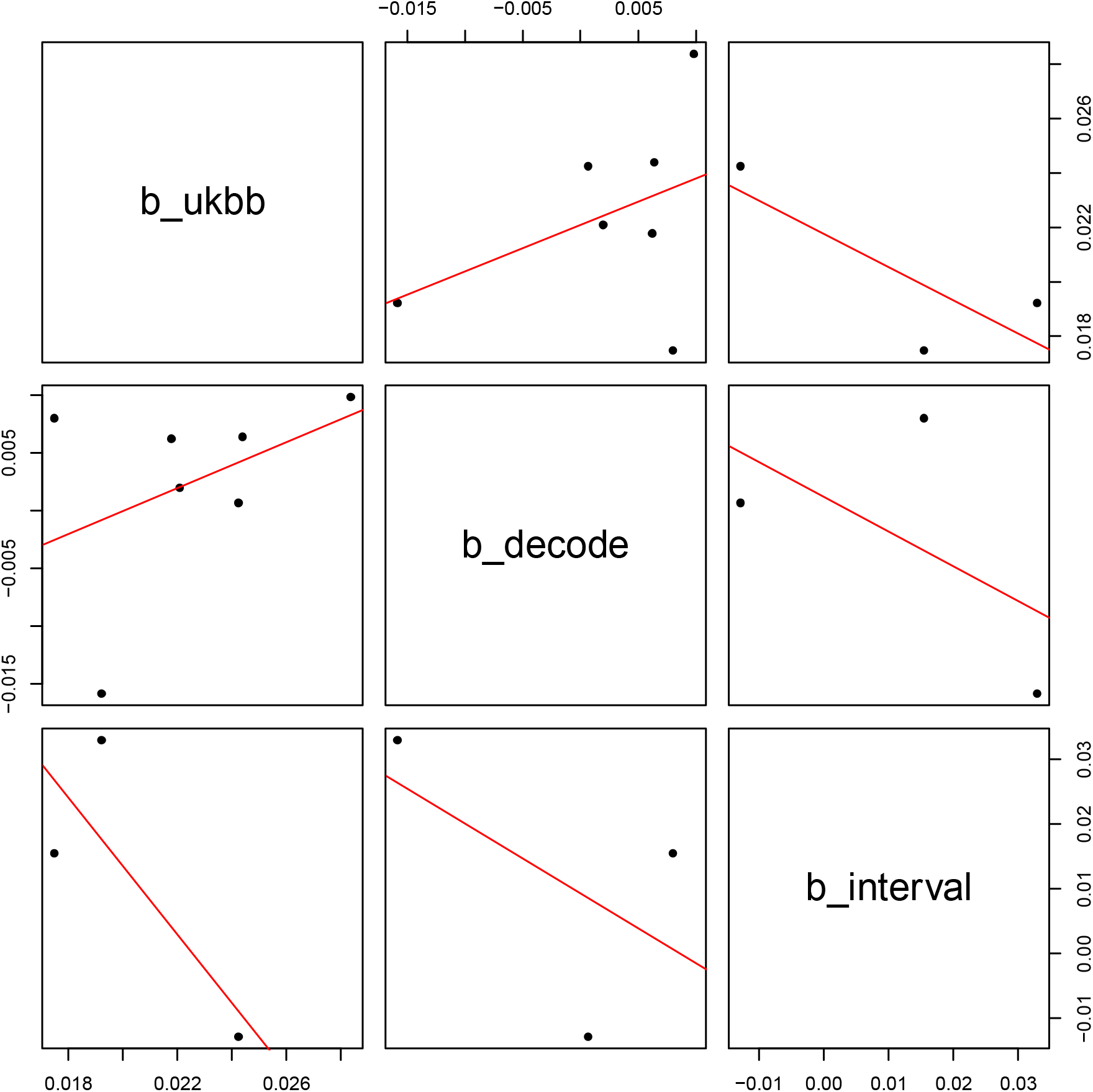
Proteins associated with non-glioblastoma PRS in the UKBB cohort compared with these proteins in deCODE and INTERVAL. There were no proteins in the UKBB cohort associated with non-glioblastoma PRS that were measured in the SCALLOP cohort.

### Protein enrichment analysis

Next, we used STRINGDB to see what processes are linked with proteins that are associated with a high genetic liability to glioma entities. We also wanted to see if the proteins associated with a high genetic liability to glioma entities were linked more than expected by chance, so we calculated an empirical p-value for the enrichment of these proteins. We were unable to include BAGE3 and protein isoform LL of CAMP in this analysis as they were not available in STRINGDB.

We found a proportion of proteins associated with the glioma PRS to be associated with the keywords signal, glycoprotein, and disulfide bond. The gene ontology (GO) processes linked to proteins associated with the glioma PRS included response to external stimulus, immune system process, and defense response (Supplementary Table 11). STRINGDB enrichment for the 160 proteins discovered in the glioma analysis is <1.06×10-16 (Figure 5), and the empirical p-value is <0.05 which suggests the enrichment is more than expected by chance.

**Figure 5:**
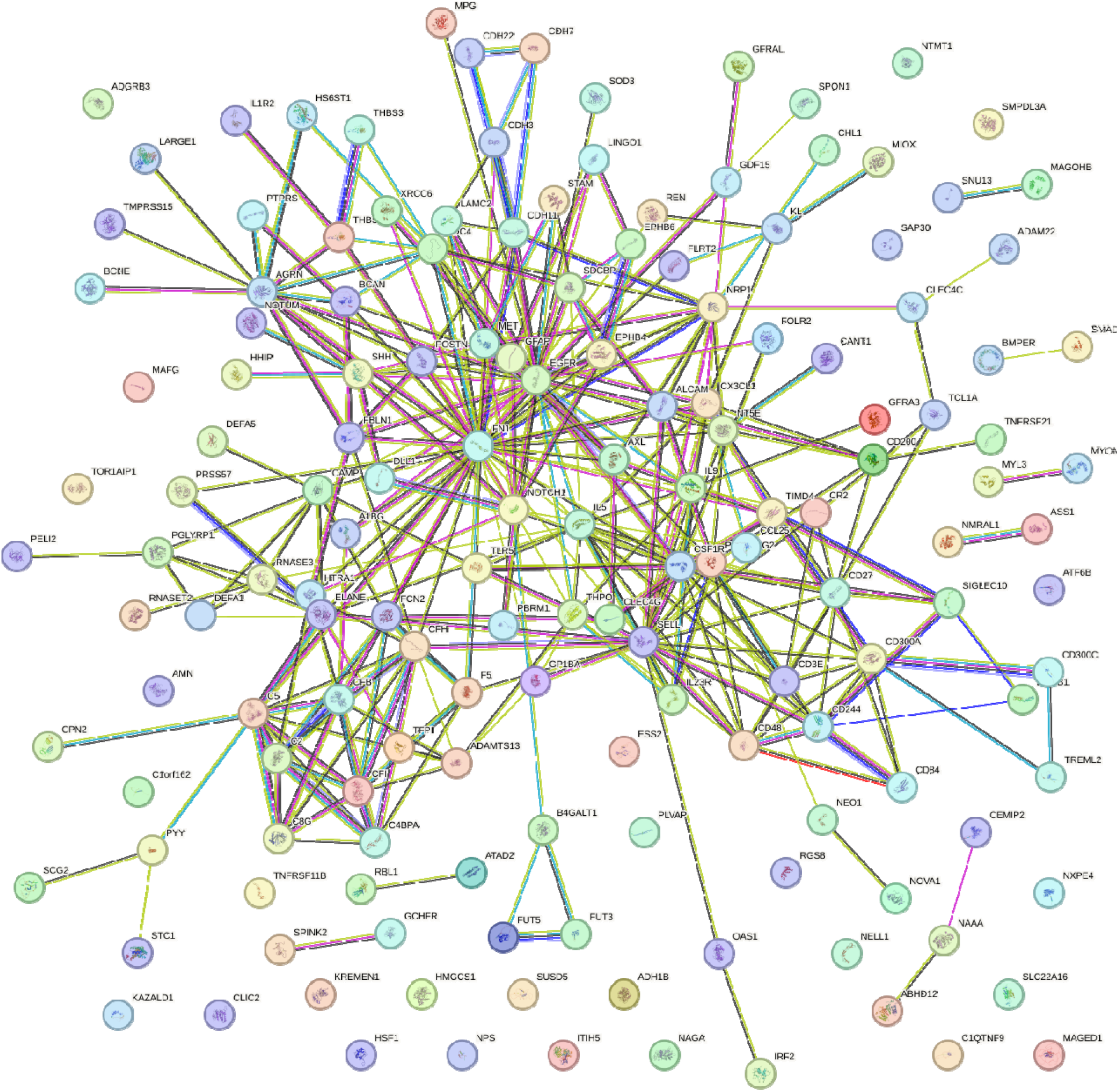
STRINGDB evidence of co-expression in proteins associated with glioma PRS.

We identified a number of proteins linked with the glioblastoma PRS to be associated with the keywords, signal, disulfide bond, and glycoprotein. The GO processes associated with the proteins linked to glioblastoma PRS were immune system process, cell adhesion, and defense response (Supplementary Table 12). STRINGDB enrichment for the 78 proteins discovered in the glioblastoma analysis is 7.26×10-13 (Figure 6), and the empirical p-value<0.01 which suggests the enrichment is more than expected by chance.

**Figure 6:**
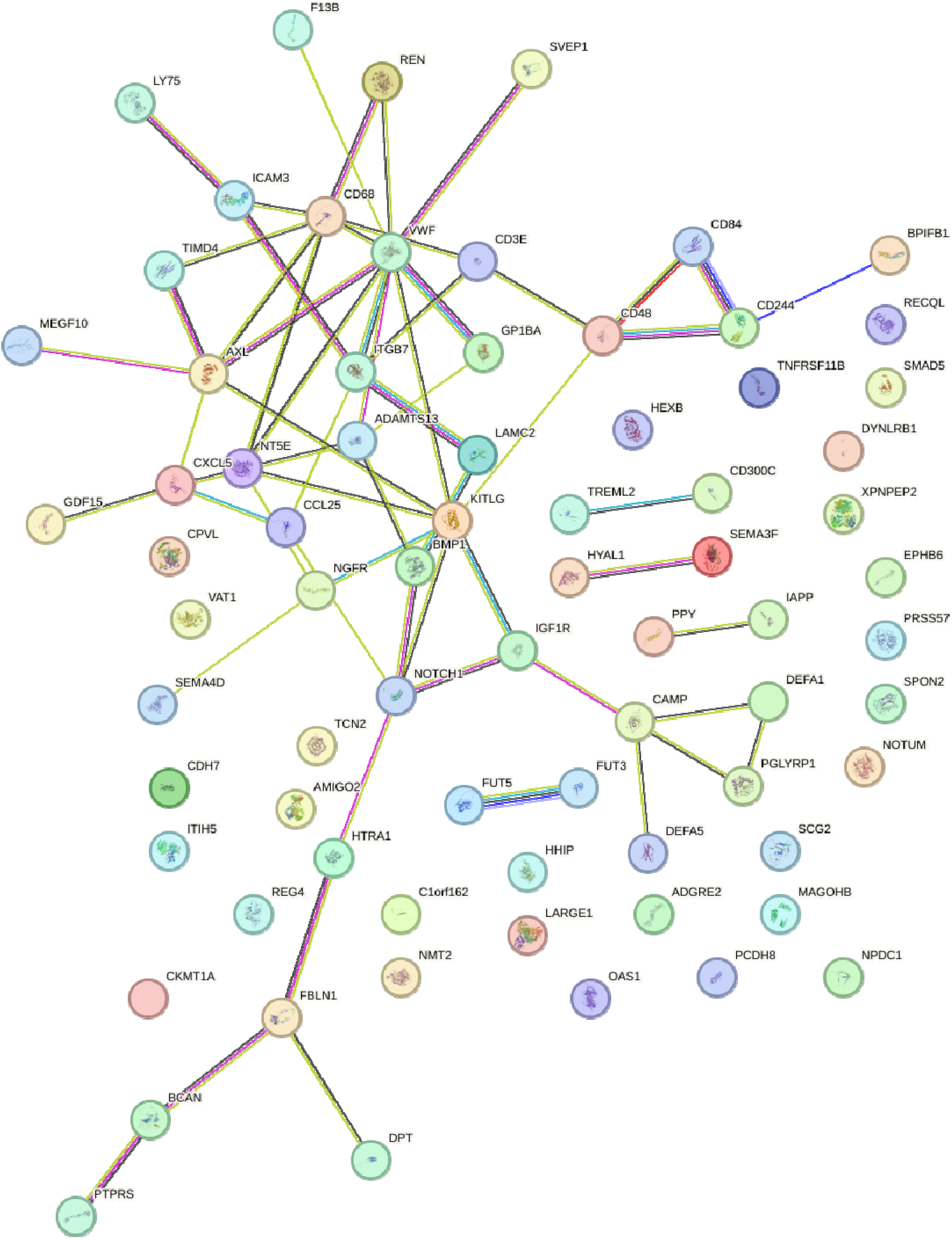
STRINGDB evidence of co-expression in proteins associated with glioblastoma PRS.

We found 10 of the proteins associated with the non-glioblastoma PRS to be linked with the term signal, and 6 of the proteins to be linked with the GO component cell surface (Supplementary Table 13). STRINGDB enrichment for the 13 proteins discovered in the non-glioblastoma analysis is 0.4803, and the empirical p-value was found to be 0.5 which suggests the enrichment could be due to chance.

### Forward MR

There were 13 associations found in the forward MR analysis. The strongest association of these was genetically-proxied circulating pleckstrin homology like domain family B member 1 (PHLDB1) and non-glioblastoma risk (Table 2). Genetically-proxied circulating PHLDB1, electron transfer flavoprotein subunit alpha (ETFA), ribonucleoside-diphosphate reductase large subunit (RIR1) were found to have a strong association, hemoglobin subunit zeta (HBZ) and acetylcholinesterase (ACHE) were found to have a suggestive association with glioma risk. Genetically-proxied circulating RIR1 was the only protein to have strong evidence, and genetically-proxied circulating transcription elongation factor A2 (TCEA2) had suggestive association with glioblastoma risk. Genetically-proxied circulating PHLBD1, ETFA, RAC-gamma serine/threonine kinase (AKT3), trehalase (TREH), butyrophilin subfamily 3 member A1 (BT3A1) were found to have a strong association, and angiopoietin-related protein 3 (ANGPTL3) has a suggestive association with non-glioblastoma risk. No associations between glioma, glioblastoma and non-glioblastoma were identified in the INTERVAL and SCALLOP cohorts.

**Table 2:** Associations between glioma entities and proteins in UKBB cohort in the forward Mendelian randomization analysis.

| Protein Dataset | Glioma Entity | Protein | Method | Number of SNPs | Effect (95% Confidence Interval) | Q-value |
| --- | --- | --- | --- | --- | --- | --- |
| UKBB | Non-glioblastoma | PHLDB1 | Wald ratio | 1 | -6.257 (-7.739 to -4.775) | 2.40x10<sup>-13</sup> |
|  | Non-glioblastoma | AKT3 | Wald ratio | 1 | -2.303 (-3.177 to -1.428) | 2.33x10<sup>-4</sup> |
|  | Glioma | PHLDB1 | Wald ratio | 1 | -2.770 (-3.889 to -1.652) | 2.29x10<sup>-3</sup> |
|  | Non-glioblastoma | TREH | Inverse variance weighted | 5 | -0.364 (-0.520 to -0.208) | 2.94x10<sup>-3</sup> |
|  | Glioma | HBZ | Inverse variance weighted | 5 | -0.150 (-0.226 to -0.074) | 6.95x10<sup>-2</sup> |
|  | Glioma | ACHE | Inverse variance weighted | 4 | 0.247 (0.122 to 0.372) | 6.95x10<sup>-2</sup> |
|  | Non-glioblastoma | ANGPTL3 | Inverse variance weighted | 3 | 0.325 (0.155 to 0.494) | 8.47x10<sup>-2</sup> |
| deCODE | Non-glioblastoma | ETFA | Wald ratio | 1 | 3.304 (2.394 to 4.214) | 1.77x10<sup>-9</sup> |
|  | Glioma | ETFA | Wald ratio | 1 | 1.724 (1.027 to 2.421) | 1.95x10<sup>-3</sup> |
|  | Glioma | RIR1 | Wald ratio | 1 | 1.421 (0.756 to 2.086) | 2.23x10<sup>-2</sup> |
|  | Non-glioblastoma | BT3A1 | Wald ratio | 1 | 2.151 (1.139 to 3.163) | 2.43x10<sup>-2</sup> |
|  | Glioblastoma | RIR1 | Wald ratio | 1 | 1.699 (0.900 to 2.498) | 4.81x10<sup>-2</sup> |
|  | Glioblastoma | TCEA2 | Wald ratio | 1 | -0.644 (-0.969 to -0.318) | 8.17x10<sup>-2</sup> |

In our colocalization analysis, associations with ETFA, RIR1 and BT3A1 had a PPH4>80% suggesting strong evidence of colocalization. AKT3 and non-glioblastoma PPH4=60.08% suggesting there is moderate evidence of colocalization and a variant is shared. However, associations with PHLDB1, TREH and ANGPTL3 were found to have a PPH2>80% which suggests causal variant is linked with the glioma entity dataset, and associations with ACHE had a PPH1>80% suggesting the causal variant is linked with the protein dataset. Associations with HBZ and TCEA2 had a PPH3>90% suggesting the variants are distinct in the protein and glioma datasets and are in LD. Steiger filtering suggested ETFA, RIR1, ACHE, HBZ and glioma risk, TCEA2, RIR1 and glioblastoma risk, BT3A1, AKT, ANGPTL3, TREH and non-glioblastoma risk were in the correct direction of exposure to outcome.

### Protein enrichment analysis

As there were fewer than 10 proteins associated with each glioma entity in the forward MR, for the protein enrichment analysis we used results which reached genome-wide significance from the glioma GWAS(19) in which the PRS was derived to investigate potential pathways which may be implicated on the causal path.

For the glioma GWAS, the following pathways were enriched; glioblastoma signaling pathways, head and neck squamous cell carcinoma, and regulation of G1/S transition of mitotic cell cycle. These same pathways were also found to be enriched in the glioblastoma GWAS. The non-glioblastoma GWAS were found to have glioblastoma signaling pathways, cell cycle, and oncogene induced senescence.

### Observational data

In our literature search, two proteins identified in our reverse MR analysis have been reported as pre-diagnostic markers for glioma entities. Elevated levels of EGFR were also identified as a pre-diagnostic marker for glioblastoma (OR=1.58, 95% CI=1.13-2.22)(33). Increased levels of GFAP were reported as a marker for brain cancer pre-diagnosis using the UKBB cohort (Hazard ratio=1.56, 95% CI=1.31-1.86)(34).

## Discussion

In this study, we aimed to see if reverse MR could identify non-causal biomarkers which may predict disease. We also carried out forward MR analyses to identify potential markers associated with the risk of glioma to ensure the proteins identified in the reverse MR were not on the causal pathway. We also looked at observational data of protein levels before glioma diagnosis to see if this followed the same trend as our results.

Overall, we identified 161 proteins to be associated with the glioma PRS, 79 proteins with the glioblastoma PRS, and 13 proteins with non-glioblastoma PRS. Enrichment analyses identified a proportion of plasma proteins to be associated with the PRS of glioma and glioblastoma to be correlated with response to external stimulus and immune response. A nested case-control study investigated immunoglobulin E (IgE) levels before glioma diagnosis, which found risk of glioma to be inversely related to allergic sensitization, interestingly this was more notable in women(35). A similar nested case-control study also identified a positive total IgE (>100kU/L) was associated with decreased risk of glioma in both sexes, this was evidenced at least 20 years before diagnosis(36). For glioblastoma, this association was only identified in women with an allergen-specific IgE (>0.35kU_A_ /L)(36). These two studies find IgE levels to decrease risk of glioma, our study finds similar results as we suggest immune system processes to be an early marker for glioma. We did not find any of the suggested proteins found in the forward MR to be associated with any of the proteins discovered in the reverse MR, suggesting these are non-causal.

In our forward MR analyses we suggest 10 unique proteins to be associated with risk of glioma entities. The associations which passed forward MR analyses and colocalization was genetically-proxied circulating ETFA and RIR1 in glioma risk, RIR1 in glioblastoma risk, and ETFA, BT3A1, AKT3 in non-glioblastoma risk. AKT3 has previously been associated with the development of IDH-mutant gliomas, with higher levels of mRNA in lower grade gliomas(37, 38). PHLDB1, TREH, TCEA2 and ETFA have also previously been identified as risk loci (19, 39, 40). We identified 5 putative proteins, RIR1, BT3A1 HBZ, ACHE, ANGPTL3, which have not been identified in glioma risk. However, HBZ, ACHE, and ANGPTL3 were not suggested to colocalize.

We investigated the pathways involved in markers which reached genome-wide significance in the GWAS which the PRS of glioma entities were derived from(19). In this analysis glioma and glioblastoma was associated with glioblastoma signaling pathways, these include DNA repair, G1/S progression, cell cycle progression, and cell migration(41). This differs from the pathways found to be enriched in our reverse MR analysis which includes response to external stimulus and immune system process(41). This provides more evidence to our hypothesis that reverse MR may detect early markers for disease.

We searched the literature for proteins which were assessed pre-diagnostically of glioma. Our search identified EGFR and GFAP(33, 34). Increased circulating levels of EGFR was associated with glioblastoma risk(33), and increased levels of GFAP were associated with brain cancer risk(34), both of which were assessed before diagnosis. Similarly, in our study we identified reverse MR associations usually found increased circulating EGFR and GFAP in reverse MR associated with glioma, glioblastoma and non-glioblastoma analysis compared to the forward MR analysis (Table 3-4). This association differed in the forward MR which suggested these circulating markers would be decreased.

**Table 3:** Associations between glioma entities and EGFR and GFAP in the forward Mendelian randomization analysis.

| Dataset | Glioma Entity | Protein | Method | Number of SNPs | Effect (95% Confidence Interval) | P-value |
| --- | --- | --- | --- | --- | --- | --- |
| UKBB | Glioblastoma | EGFR | Inverse variance weighted | 3 | -0.064 (-0.318 to 0.190) | 0.620 |
| deCODE | Glioblastoma |  | Inverse variance weighted | 3 | -0.229 (-0.568 to 0.110) | 0.185 |
| UKBB | Glioma |  | Inverse variance weighted | 3 | -0.031 (-0.283 to 0.221) | 0.807 |
| deCODE | Glioma |  | Inverse variance weighted | 3 | -0.226 (-0.575 to 0.123) | 0.205 |
| UKBB | Non-Glioblastoma |  | Inverse variance weighted | 3 | 0.097 (-0.219 to 0.412) | 0.549 |
| deCODE | Non-Glioblastoma |  | Inverse variance weighted | 3 | -0.121 (-0.467 to 0.225) | 0.493 |
| UKBB | Glioma | GFAP | Inverse variance weighted | 4 | -0.030 (-0.350 to 0.290) | 0.856 |
| UKBB | Glioblastoma |  | Inverse variance weighted | 4 | -0.092 (-0.482 to 0.298) | 0.644 |
| UKBB | Non-Glioblastoma |  | Inverse variance weighted | 4 | 0.048 (-0.371 to 0.466) | 0.823 |

**Table 4:** Associations between glioma entities and EGFR and GFAP in the reverse Mendelian randomization analysis.

| Dataset | Protein | Glioma Entity | Method | Number of SNPs | Effect (95% Confidence Interval) | P-value |
| --- | --- | --- | --- | --- | --- | --- |
| UKBB | EGFR | Glioblastoma | Inverse variance weighted | 85 | -0.002 (-0.016 to 0.011) | 0.725 |
| deCODE |  | Glioblastoma | Inverse variance weighted | 83 | -0.006 (-0.019 to 0.007) | 0.367 |
| INTERVAL |  | Glioblastoma | Inverse variance weighted | 87 | 0.038 (0.002 to 0.074) | 0.039 |
| UKBB |  | Glioma | Inverse variance weighted | 99 | 0.011 (-0.003 to 0.025) | 0.110 |
| deCODE | | Glioma | Inverse variance weighted | 99 | -0.026 (-0.040 to -0.011) | $7.45 \times 10^{-4}$ |
| INTERVAL |  | Glioma | Inverse variance weighted | 99 | 0.012 (-0.031 to 0.054) | 0.592 |
| UKBB | | Non-Glioblastoma | Inverse variance weighted | 91 | 0.019 (0.009 to 0.030) | $3.20 \times 10^{-4}$ |
| deCODE | | Non-Glioblastoma | Inverse variance weighted | 92 | -0.016 (-0.027 to -0.004) | $7.13 \times 10^{-3}$ |
| INTERVAL |  | Non-Glioblastoma | Inverse variance weighted | 93 | 0.033 (-0.002 to 0.068) | 0.064 |
| deCODE | GFAP | Glioblastoma | Inverse variance weighted | 83 | 0.005 (-0.016 to 0.011) | 0.427 |
| UKBB | | Glioblastoma | Inverse variance weighted | 85 | 0.020 (0.006 to 0.033) | $5.93 \times 10^{-3}$ |
| deCODE |  | Glioma | Inverse variance weighted | 99 | 0.008 (-0.005 to 0.020) | 0.233 |
| UKBB | | Glioma | Inverse variance weighted | 99 | 0.030 (0.016 to 0.045) | $4.75 \times 10^{-5}$ |
| deCODE |  | Non-Glioblastoma | Inverse variance weighted | 92 | 0.005 (-0.005 to 0.016) | 0.298 |
| UKBB | | Non-Glioblastoma | Inverse variance weighted | 91 | 0.015 (0.004 to 0.026) | $8.45 \times 10^{-3}$ |

Although there were many advantages to our study, there were also some limitations. The glioma GWAS cohort was split into glioblastoma (6,191 cases) and non-glioblastoma (5,819), which meant in the all glioma cases analysis, there could be a slight bias towards glioblastoma compared to non-glioblastoma patients. There is also a potential source of more bias in our results as we combined heterogenous tumours together (glioblastoma and non-glioblastoma) within the all glioma cases cohort, this makes it difficult to find appropriate associations in all glioma cases. This potential bias is also true for the non-glioblastoma cohort where glioma cases not classified as glioblastoma are put into the same heterogenous group. For future glioma studies, larger sample sizes and independent subtype analyses may be required to achieve more statistical power and reduce potential bias.

We were also limited to sample size of the pQTL dataset. Only recently have large blood pQTL datasets been published which assay thousands of proteins, however these GWAS are limited to fewer individuals compared to eQTL studies (7–10). This leads to some results lacking statistical power as well as making it harder to replicate our findings. The deCODE study included individuals from the Icelandic cancer project (∼52%) diagnosis with some individuals already diagnosed with cancer, although the size of the population included the chances are only one individual may be diagnosed(21, 22). There is a further issue of transportability of our results, as we only include individuals of European descent these findings may not be generalizable to different populations or individuals of other ethnic backgrounds. There is also an issue of temporality, protein levels are measured from the general population, most of whom will not have glioma or go on to develop glioma.

Our criteria for each association reaching q-value<0.05 for P1 and an MR P-value<0.05 for P4 may have excluded other potential biomarkers which may be associated with genetic liability for glioma. The reverse MR and forward MR also assumes no pleiotropy within the analysis, which is also known as the exclusion restriction criterion. Horizontal pleiotropy is when a SNP influencing both the glioma liability and circulating protein by independent routes. Vertical pleiotropy occurs due to a SNP influencing other traits which has an impact on circulating protein levels via the effect on glioma liability(42, 43). As we have a PRS approach, this analysis is less likely to have pleiotropic effects as we are looking at the effect of multiple SNPs. To further limit the potential effects of horizontal pleiotropy the glioma GWAS was clumped at an LD threshold of r^2^=0.1. However, in our forward MR approach horizontal pleiotropy can be a concern. For associations that are driven by a single SNP it was not possible to carry out MR sensitivity analyses, or heterogeneity tests. These associations can also be prone to horizontal pleiotropy and issues with statistical power.

It is also important to note that our analysis presents reverse MR results, suggesting these putative proteins are associated with genetic liability to glioma. There has not been a prospective clinical study to provide more evidence of this association. Further studies will need to be carried out to delineate the association of the potentially predictive proteins as diagnostic blood biomarkers for glioma, as well as further research into understanding the role of these proteins in the context of glioma.

## Conclusion

Our findings identify a high genetic liability to glioma is associated with the immune system processes. Non-causal plasma biomarkers identified through PRS associations could indicate novel biomarkers of early glioma development. Candidate markers, including ETFA, RIR1 and BT3A1 were associated with risk of glioma entities using forward MR. Enrichment analyses suggest different pathways are enriched in potentially causal versus non-causal markers.

## Data Availability

Data available in the supplementary materials.

## Conflicts of Interest

JWR is employed at Boehringer Ingelheim for unrelated research. The other authors have no conflicts of interest to declare.

## Author Contributions

GH and KMK conceived the study. GH and LJA designed the study. LJA performed all statistical analyses. LJA drafted the manuscript. GH, JWR and KMK supervised the project. GH and JWR provided critical manuscript revisions. All authors read and reviewed the final manuscript.

## Notes

### Funding Statement

LJA and KMK are funded by Cancer Research UK (grant number C30758/A29791); ZAT is funded by Southmead Hospital Charitable Funds: Brain tumour bank and research fund 8036. KMK is funded by Innovate [grant number 10027624]. GH is supported by the Wellcome Trust and Royal Society [208806/Z17Z] and the Medical Research Council (MRC) Integrative Epidemiology Integrative Epidemiology Unit (IEU) at the University of Bristol [MC_UU_00032/01]. This work was supported by Cancer Research UK [grant number C18281/A29019 and C18281/A30905].

