## Supplementary Information for "Reverse Mendelian randomization separates causes from early proteomic biomarkers of glioma"

Method:

We carried out a heterogeneity analysis by performing a Cochrane’s Q-test in associations with 2 or more SNPs. If the Cochrane’s Q-test<0.05 this result would be deemed as potential heterogeneity. As before, if an association had potential heterogeneity, we carried out a leave one out analysis, if the SNPs differed by >0.01 this was deemed as a highly heterogenous association. If the forward MR included 3 or more SNPs in the association, we were also able to look at the direction of effect in in MR-Egger and weighted median regression scores to ensure these were in the same direction as the MR result.

Result:

All associations with 2 or more SNPs were found to have the same direction of effect across MR-Egger and weighted median regression. These associations were also tested for potential heterogeneity. Each association had a Cochrane’s q-test>0.05, except for the association between non-glioblastoma and TREH which did not pass this threshold or the leave one out analysis.

Supplementary section 1: Heterogeneity test and MR effect estimate sensitivity analyses.
